# Aspergillus and Candida in primary and secondary immunodeficiencies in USIDNET

**DOI:** 10.64898/2026.09.11.26362658

**Authors:** Samantha Y. Starkey, USIDNET Consortium, Kathleen E Sullivan

## Abstract

**Objective:** Fungal infections can represent opportunistic infections leading to the diagnosis of immunocompromise. Specific inborn errors of immunity are associated with aspergillus but it is unclear how sensitive aspergillus is for the subsequent diagnosis of those inborn errors of immunity.

**Methods:** The patient registry USIDNET was utilized to extract clinical conditions and laboratory values from patients with candida and aspergillus diagnosis codes. We restricted the diagnoses to deep infections and then identified the associated clinical conditions along with potential settings, such as bone marrow transplant.

**Results:** We identified 41 patients with aspergillus as a diagnosis code. The types of diagnoses associated with aspergillus included T cell disorders, antibody disorders, as well as the anticipated myeloid disorders. Bone marrow transplant was identified as a potential predisposing condition. 64. Patients were identified with candida as a diagnosis code. Similarly, T cell disorders, antibody disorders, and myeloid disorders were found.

**Conclusions:** Candida and aspergillus are generally considered sentinel infections for myeloid disorders and T cell disorders. However, we found these infections associated with a broad range of inborn errors of immunity. This suggests that diagnostic testing should be broadened when facing patients with these infections.

---

To the Editor

*Aspergillus* and *Candida* cause opportunistic infections that occur primarily in immunocompromised patients. *Aspergillus* conidia (spores) are ubiquitous in the environment and clinical manifestations of disease after conidia inhalation is dependent upon host immune response, ranging from allergic bronchopulmonary aspergillosis and asthma with fungal sensitization to pulmonary aspergillosis and invasive aspergillosis [1]. Important pathways in the host response to *aspergillus* include the nicotinamide adenine dinucleotide phosphate (NADPH)-dependent reactive oxidative species production which is impaired in chronic granulomatous disease (CGD) [1]. Recipients of hematopoietic stem cell transplant (HCT), solid organ transplant, and immunosuppressive therapies for rheumatologic diseases and malignancies are also at higher risk of invasive aspergillosis [1]. Susceptibility to *Aspergillus* is complex with contributions from neutrophils, T cells and complement [1]. In contrast, *Candida* species are part of the normal skin and mucosal microflora. Clinical manifestations include mucocutaneous and invasive disease. Adaptive immunity is thought to primarily control mucosal candida infection by interleukin-17 (IL-17)–producing lymphocytes, while the innate immune system is critical for preventing systemic candidiasis [2].

Both *Aspergillus* and invasive *Candida* are sentinel infections and felt to indicate inborn errors of immunity (IEI). To understand the range of immunodeficiency states in which *Aspergillus* and *Candida* are seen, we utilized the United States Immunodeficiency Network (USIDNET). USIDNET collects all diagnoses, medications, and demographic information on patients with primary immunodeficiencies across participating institutions in the United States. Because it can be difficult to determine if an immunodeficiency is primary or secondary, there may be some cases of suspected secondary immunodeficiencies in the registry. Patient data is deidentified, but longitudinal data is traceable through unique identifiers.

## Methods

We performed a retrospective analysis of patients with *Aspergillus*- or *Candida*-related International Classification of Disease (ICD) diagnosis codes reported in the USIDNET registry. For *Candida*, we included only esophagitis, endocarditis, pulmonary candidiasis, laryngitis, sepsis, meningitis, and vulvovaginitis (excluding conditions such as candidal diaper rash which are less unique to the immunocompromised population). The search was run on February 24th, 2026. IEI were classified based on available data according to the 2024 update on the classification from the International Union of Immunological Societies Expert Committee [3].

## Results

41 subjects in USIDNET were identified as having *Aspergillus* as a diagnosis code (Table 1 and Supplemental Table 1). Thirteen individuals had immunodeficiencies affecting cellular and humoral immunity, which included five with severe combined immunodeficiency (SCID), five with combined immunodeficiency (CID), two with activated PI3K delta syndrome (APDS), and one with candidiasis-endocrinopathy syndrome. Twelve people had predominantly antibody deficits, including six with combined variable immunodeficiency (CVID), one of which had X-linked agammaglobulinemia, and the remaining six with hypogammaglobulinemia/immunoglobulin deficiency. Fourteen individuals had congenital defects of phagocyte number or function including 13 with CGD and one with GATA2 related myelodysplasia. The patient with GATA2 related myelodysplasia and eight of the individuals with CGD underwent HCT. One individual had dyskeratosis congenita and a lung transplant. The remaining individual did not have an IEI in their diagnosis code but was on systemic corticosteroids. Overall, ten of the patients were deceased at the time of data extraction. Of the deceased individuals, two had underlying immunodeficiencies with an anticipated shortened lifespans: ataxia telangiectasia and dyskeratosis congenita, three were recipients of a solid organ transplant (two lung and one heart transplant), and three were HCT recipients.

**Table 1:** Inborn errors of immunity diagnoses in individuals with *Aspergillus* and invasive *Candida* infections in the USIDNET registry. Abbreviations: APECED: autoimmune polyendocrinopathy-candidiasis-ectodermal dystrophy; CGD: chronic granulomatous disease; CTLA4: cytotoxic T-lymphocyte–associated protein 4; CVID: common variable immunodeficiency; STAT1: signal transducer and activator of transcription 1

| Diagnoses | *Aspergillus* infections<br>(n=41) | Invasive *Candida* infections<br>(n=64) |
| --- | --- | --- |
| Immunodeficiencies affecting cellular and humoral immunity (e.g. SCID) | 13 | 15 |
| Combined immunodeficiency with associated or syndromic features (e.g. DiGeorge Syndrome, Hyper IgE) |  | 13 |
| Predominantly antibody disorders (e.g. CVID, hypogammaglobulinemia) | 12 | 16 |
| Congenital defects of phagocyte number or function (e.g. CGD) | 14 | 3 |
| Bone marrow failure (dyskeratosis congenita) | 1 |  |
| Diseases of Immune Dysregulation (e.g. APECED, CTLA4) |  | 3 |
| Defects in intrinsic and innate immunity (e.g. STAT1) |  | 5 |
| Not specified | 1 | 9 |

64 patients in USIDNET had *Candidal* infections of the following types (mutually inclusive as some patients had more than one): esophagitis (n=46), sepsis (n=14), vulvovaginitis (n=5), pulmonary candidal infection (n=1), laryngitis (n=1), and/or meningitis (n=1). One patient had chronic mucocutaneous candidiasis. Supplemental table 2 lists the patients’ associated conditions. Fifteen individuals had immunodeficiencies affecting cellular and humoral immunity including four with SCID, ten with CID/Other. CID/Other diagnoses included LRBA deficiency (n=1), Autoimmune Polyendocrinopathy-Candidiasis-Ectodermal Dystrophy (APECED) syndrome (n=1), WHIM syndrome (n=1), MHC deficiency (n=1), and multidysplastic syndrome (MDS)/GATA2 (n=1). Six of these fifteen patients received HCT. Thirteen patients had CID with associated or syndromic features: DiGeorge syndrome (n=9), HyperIgE (n=2), Kabuki syndrome (n=1), and ectodermal dysplasia (n=1). The latter individual was an HCT recipient. Sixteen individuals had primarily antibody disorders: CVID (n=7, including one with NFKB2 deficiency), hypogammaglobulinemia (n=5), hyper IgM (n=2), PTEN deficiency (n=1), and/or unspecified primary antibody disorder (n=1). Three individuals had diseases of immune dysregulation: CTLA4 (n=2) and APECED (n=1). Three individuals had CGD; all three had *Candida* sepsis and two were deceased. Five patients had STAT1 mutations (unspecified). Specific IEI diagnoses could not be determined in nine cases.

## Discussion

This study used USIDNET records to describe the range of immunodeficiencies in which *Aspergillus* and *Candida* infections may occur. In many instances, fungal infections developed in individuals with several risk factors including IEI diagnoses and immunosuppressive therapies. Thirteen out of forty-one patients with *Aspergillus* had a diagnosis of CGD. The relationship between *Aspergillus* infections and CGD is well-described. However, there were many other primary and secondary immune deficiencies recorded amongst patients diagnosed with *Aspergillus*, including combined immunodeficiencies and medically-induced immunocompromising states. *Aspergillus* infections are being increasingly recognized in the population with secondary immunosuppression due to increasing use of immunotherapeutic agents [1]. Indeed, aspergillosis is the most common invasive fungal infection among recipients of HCT [4] In our cohort, some patients with IEI also received immunosuppressive therapies and transplants that likely further increased their susceptibility to aspergillus infection.

The *Candida* cohort included twenty-eight patients with combined cellular and humoral immunodeficiencies; with the majority of them having *Candida* esophagitis (n=16), vulvovaginitis (n=3), and laryngitis (n=1); consistent with prior literature that T-cell immunodeficiency is a risk factor for mucocutaneous candidal infection [5]. Mucocutaneous infection was also seen in IEI not characterized by severe T cell dysfunction including STAT1 gain-of-function (n=3; all with esophagitis). Gain-of-function STAT1 mutations were seen in the *Candida* cohort and not the *Aspergillus* cohort, consistent with prior work indicating the role of IL-17 in immunity against mucosal candidal infections [5]. Prior work has shown two distinct pathways in neutrophil killing mechanisms for *Candida*, one of which was independent of NADPH-dependent reactive oxygen species creation [2], which is perhaps why CGD was more prevalent in the *Aspergillus* cohort than the *Candida* cohort. In the *Candida* cohort, the three patients with CGD all had *Candida* sepsis, and one also had *Candida* meningitis.

There were some patients in both cohorts where medical immunocompromise was the probable risk factor for infection. Common immunosuppressants in our cohort included corticosteroids which suppress the innate immune response to fungal pathogens and calcineurin inhibitors which disrupt the calcium–calcineurin–NFAT (nuclear factor of activated T cells) pathway [1].

Limitations of the present study include relying on the accuracy of coding and low sample sizes, particularly in patients under ten years of age. Additionally, it could not be determined whether certain immune deficits (i.e. hypogammaglobulinemia, neutropenia, pancytopenia) were related to primary immune disorders or were consequences of immunosuppressive medication.

In summary, this study used USIDNET records to describe 41 patients with *Aspergillus* infections and 64 patients with *Candida* infection including their immunological diagnoses and key immunosuppressive therapies. Both primary and secondary immunodeficiencies were seen in these cohorts, and sometimes in the same patient.

## Data Availability

All data produced in the present study are available upon reasonable request to the authors

## Appendix

The U.S. Immunodeficiency Network (USIDNET), a program of the Immune Deficiency Foundation (IDF), is supported by a cooperative agreement, U24AI86837, from the National Institute of Allergy and Infectious Diseases (NIAID).

*USIDNET Consortium – investigators and enrolling sites that contributed to this manuscript* Vichare, Vaibhavi^1^, Kilich, Gonench, MD^2^, Xiao, Rui, PhD^1^, Cunningham-Rundles, Charlotte, MD PhD^3^, Abraham, Roshini S., PhD^4^, Puck, Jennifer, MD^5^, Fuleihan, Ramsay, MD^6^, Marsh, Rebecca, MD^7^

^1^ Department of Biomedical and Health Informatics, Children’s Hospital of Philadelphia, Philadelphia, PA 19146, USA

^2^ Division of Allergy Immunology, Children’s Hospital of Philadelphia, Philadelphia, PA 19104. USA.

^3^ Department of Medicine, Department of Pediatrics, Icahn School of Medicine at Mount Sinai New York City NY, 10029

^4^ Department of Pathology and Laboratory Medicine, Nationwide Children’s Hospital, Columbus, OH 43205, USA

^5^ Division of Allergy/Immunology and Blood and Marrow Transplantation, University of California San Francisco Dept. of Pediatrics, San Francisco, CA 94143 USA

^6^ Division of Allergy, Immunology and Rheumatology, Department of Pediatrics, Columbia University Irving Medical Center, New York, NY 10032, USA

^7^ Division of Bone Marrow Transplantation and Immune Deficiency, Cincinnati Children’s Hospital Medical Center, Cincinnati, Ohio; Department of Pediatrics, University of Cincinnati College of Medicine, Cincinnati, Ohio.

