## Supplemental Tables for "Aspergillus and Candida in primary and secondary immunodeficiencies in USIDNET"

*Journal of Clinical Immunology*

^1^ Nemours Children’s Hospital – Delaware, Wilmington, DE 19803 USA

^2^ Division of Allergy Immunology, Children’s Hospital of Philadelphia, Philadelphia, PA 19104 USA

* Listed in appendix

Corresponding Author

Kathleen E. Sullivan

3615 Civic Center Blvd. Philadelphia, PA 19104 USA

215-590-1697

**Supplemental Table 1: Diagnoses and medical immunosuppression in patients with *Aspergillus* infections in USIDNET**

|  | ID | Diagnosis | Medical Immunosuppression | Opportunistic Infections | Alive/Deceased |
| --- | --- | --- | --- | --- | --- |
| Immunodeficiencies affecting cellular and humoral immunity | | | | | |
| 1 | 3319 | SCID | HCT | BK, EBV, rhizopus, adenovirus viremia | Deceased |
| 2 | 1244 | SCID | HCT, kidney transplant | CMV, strep viridans | Alive |
| 3 | 4711 | SCID, Hyper IgE syndrome | | Histoplasmosis, PJP | Deceased |
| 4 | 2343 | SCID |  |  | Alive |
| 5 | 6280 | ADA-SCID |  | candida, pseudomonas | Alive |
| 6 | 687 | CID (Artemis deficiency) | HCT |  | Alive |
| 7 | 2821 | CID | Chemotherapy | pseudomonas, CMV, cryptosporidiosis, actinomycosis, candida (unspecified) | Alive |
| 8 | 2504 | CID, Ataxia telangiectasia | Chemotherapy | CMV, EBV, pseudomonas, candida (unspecified) | Deceased |
| 9 | 3627 | CID, WHIM | Splenectomy | Candida | Alive |
| 10 | 4277 | CID | HCT | Adenovirus viremia, pseudomonas | Deceased |
| 11 | 4603 | APDS, ALPS |  | CMV | Alive |
| 12 | 5254 | APDS | HCT |  | Alive |
| 13 | 4716 | Candidiasis-endocrinopathy syndrome, STAT1 | HCT | CMC, PJP | Deceased |
| Predominantly antibody disorders | | | | | |
| 1 | 3277 | CVID | HCT | BK, cryptosporidia, PJP, varicella pneumonia | Deceased |
| 2 | 2372 | CVID | Heart transplant | EBV | Alive |
| 3 | 3582 | CVID |  | Pseudomonas | Alive |
| 4 | 3603 | CVID |  |  | Alive |
| 5 | 4436 | CVID, XLA | Lung transplant | CMV, mycobacterium kansasii | Alive |
| 6 | 5397 | CVID | Steroids, calcineurin inhibitor, antimetabolite | PJP, pneumocystis, candida (unspecified) | Alive |
| 7 | 5635 | Hypogammaglobulinemia | Steroids | Mycobacterium | Alive |
| 8 | 847 | Hypogammaglobulinemia | Lung transplant | BK, candida, HSV, MAC | Alive |
| 9 | 2421 | Hypogammaglobulinemia | Heart transplant | Pseudomonas, CMV | Deceased |
| 10 | 1401 | Hypogammaglobulinemia | Steroids | Candida | Alive |
| 11 | 3575 | Hypogammaglobulinemia | | Mycobacterium avium, pseudomonas | Alive |
| 12 | 4245 | Immunoglobulin deficiency | | Pseudomonas | Alive |
| Congenital defects of phagocyte number or function | | | | | |
| 1 | 5303 | CGD | HCT, lung transplant | VZV | Deceased |
| 2 | 5339 | CGD | HCT, liver transplant | EBV | Alive |
| 3 | 2588 | CGD | HCT | Candida (unspecified) | Alive |
| 4 | 3818 | CGD | HCT | Adenovirus viremia | Alive |
| 5 | 4598 | CGD | HCT | Mycobacterium, nocardia | Alive |
| 6 | 5297 | CGD | HCT | Enterobacter cloacae | Alive |
| 7 | 6119 | CGD | HCT |  | Alive |
| 8 | 1291 | CGD | Steroids |  | Alive |
| 9 | 3018 | CGD | Steroids |  | Alive |
| 10 | 5338 | CGD | Biologics | VZV | Alive |
| 11 | 1892 | CGD |  | Strongyloides | Alive |
| 12 | 2508 | CGD |  |  | Deceased |
| 13 | 5179 | CGD |  |  | Alive |
| 14 | 3108 | GATA2-related MDS | HCT | Mycobacterium fortuitum | Alive |
| Bone marrow failure | | | | | |
| 1 | 2476 | Dyskeratosis congenita, hypogammaglobulinemia | Lung transplant | CMV, EBV | Deceased |
| Not specified | | | | | |
| 1 | 5405 |  | Steroids | Fusarium, MAC | Alive |

Clinical features at time during encounters where patients were diagnosed with *Aspergillus* infection. “Steroids” indicates systemic corticosteroid use. “Unspecified” immunosuppression indicates that a patient had a diagnosis code for immunosuppressive medication use but the exact agent was not specified. Abbreviations: ADA: adenosine deaminase; ALPS: autoimmune lymphoproliferative syndrome; APDS: Activated PI3K delta syndrome; BK: BK polyomavirus; CGD: chronic granulomatous disease; CID: combined immunodeficiency; CMC: chronic mucocutaneous candidiasis; CMV: cytomegalovirus; CVID: common variable immunodeficiency; EBV: Epstein–Barr virus; GATA2: GATA-binding protein 2; HCT: hematopoietic stem cell transplant; HSV: herpes simplex virus; MAC: Mycobacterium avium complex; MDS: myelodysplastic syndrome; PJP: Pneumocystis jirovecii pneumonia; SCID: severe combined immunodeficiency; STAT1: signal transducer and activator of transcription 1; VZV: varicella zoster virus; WHIM: warts, hypogammaglobulinemia, infections, and myelokathexis; XLA: X linked agammaglobulinemia

**Supplemental Table 2: Diagnoses and medical immunosuppression in patients with *Candida* infections in USIDNET**

|  | ID | Diagnosis | Medical Immunosuppression | Opportunistic Infections | Alive/Deceased | Type of candidal infection |
| --- | --- | --- | --- | --- | --- | --- |
| Immunodeficiencies affecting cellular and humoral immunity | | | | | | |
| 1 | 3069 | MHC I and II deficiency, hypogammaglobulinemia | HCT, steroids, splenectomy | BK, cryptosporidiosis, CMV, herpesviral meningoencephalitis, salmonella sepsis | Deceased | sepsis |
| 2 | 1346 | SCID | HCT, intestinal transplant | cryptosoridiosis | Alive | sepsis |
| 3 | 3927 | SCID | HCT | VZV, pseudomonas | Alive | esophagitis |
| 4 | 6280 | ADA-SCID |  | aspergillus, pseudomonas | Alive | esophagitis |
| 5 | 6409 | SCID |  |  | Alive | esophagitis |
| 6 | 672 | CID |  | proteus | Alive | esophagitis |
| 7 | 2398 | CID |  |  | Alive | vaginitis |
| 8 | 1399 | CID, Cornelia de Lange |  |  | Alive | sepsis |
| 9 | 2776 | CID, LRBA deficiency | HCT | CMV pneumonitis | Alive | esophagitis |
| 10 | 2818 | CID | HCT | cryptosporidiosis, CMV viremia, EBV, Mycobacterium chelonae, PJP | Deceased | sepsis |
| 11 | 3225 | CID | HCT, biologic, steroids |  | Alive | esophagitis |
| 12 | 3425 | CID, APECED syndrome | Steroids |  | Alive | esophagitis |
| 13 | 3627 | CID, WHIM | Splenectomy | aspergillus | Alive | esophagitis |
| 14 | 6279 | CID, MDS/GATA2 |  |  | Deceased | sepsis |
| 15 | 6347 | CID | Steroids | cryptococcus, CMV, mycobacteria, pseudomonas | Deceased | esophagitis |
| Combined immunodeficiency with associated or syndromic features | | | | | | |
| 1 | 659 | HyperIgE |  |  | Alive | esophagitis |
| 2 | 4571 | HyperIgE, HIV |  | MAC | Alive | esophagitis |
| 3 | 260 | DiGeorge syndrome, hypogammaglobulinemia |  |  | Deceased | esophagitis |
| 4 | 5823 | DiGeorge syndrome, hypogammaglobulinemia | Steroids | pseudomonas | Deceased | sepsis |
| 5 | 6091 | DiGeorge syndrome, hypogammaglobulinemia |  |  | Alive | esophagitis |
| 6 | 6143 | DiGeorge syndrome, hypogammaglobulinemia |  |  | Alive | esophagitis |
| 7 | 751 | DiGeorge syndrome |  |  | Alive | esophagitis |
| 8 | 1535 | DiGeorge syndrome |  |  | Alive | laryngitis |
| 9 | 2619 | DiGeorge syndrome |  |  | Alive | vulvovaginitis |
| 10 | 6019 | DiGeorge syndrome |  | enterococcal bacteremia, pseudomonas | Alive | sepsis |
| 11 | 2756 | DiGeorge syndrome |  |  | Alive | vulvovaginitis |
| 12 | 6018 | Kabuki syndrome | Heart transplant |  | Alive | esophagitis |
| 13 | 5315 | Mutation in CFP/CYBB/IKBKG/MAGT1/XIAP, Ectodermal dysplasia | HCT, biologics, other unspecified | MAC, BK | Deceased | sepsis |
| Predominantly antibody disorders | | | | | | |
| 1 | 4518 | PTEN deficiency |  |  | Alive | esophagitis |
| 2 | 2735 | Primary antibody disorder, B-ALL, AML | HCT |  | Alive | sepsis |
| 3 | 5253 | Hyper IgM, congenital neutropenia, cyclic neutropenia, hypogammaglobulinemia | Chemotherapy |  | Alive | esophagitis |
| 4 | 1401 | Hypogammaglobulinemia | Steroids | aspergillosis | Alive | esophagitis |
| 5 | 2940 | Hypogammaglobulinemia, Monoallelic deletion of NKX2-1 gene | Lung transplant, steroids |  | Alive | vaginitis |
| 6 | 3665 | Hypogammaglobulinemia |  |  | Alive | esophagitis, vulvovaginitis |
| 7 | 5388 | Hypogammaglobulinemia |  |  | Alive | esophagitis |
| 8 | 5461 | Hypogammaglobulinemia | Nucleotide synthesis inhibitor |  | Alive | esophagitis |
| 9 | 3324 | Hyper IgM syndrome, Mutation in CFP/CYBB/IKBKG/MAGT1/XIAP, neutropenia | HCT, steroids | EBV, HSV | Alive | esophagitis |
| 10 | 193 | CVID |  |  | Alive | esophagitis |
| 11 | 2218 | CVID, B-ALL, aplastic anemia | HCT | Fungal endopthalmitis & chorioretinitis, fungal sepsis, BK | Alive | sepsis |
| 12 | 3061 | CVID |  | CMV | Alive | esophagitis |
| 13 | 3491 | CVID |  |  | Alive | esophagitis |
| 14 | 3555 | CVID |  |  | Alive | esophagitis |
| 15 | 5831 | CVID, NFKB2 deficiency | Steroids | Serratia, enterococcus sepsis | Alive | esophagitis |
| 16 | 1486 | CVID, Burkitt lymphoma, Monoallelic mutation of FLG gene | Chemotherapy, steroids | Disseminated HSV | Alive | esophagitis, sepsis |
| Diseases of immune dysregulation | | | | | | |
| 1 | 4355 | APECED syndrome |  |  | Alive | esophagitis |
| 2 | 1959 | CTLA4, ALPS | HCT |  | Alive | esophagitis |
| 3 | 6085 | CTLA4, ALPS, CVID |  |  | Alive | esophagitis |
| Congenital defects of phagocyte number or function | | | | | | |
| 1 | 1575 | CGD (X linked) | Steroids | tuberculous meningitis | Deceased | meningitis, sepsis |
| 2 | 5910 | CGD (X linked) | Unspecified |  | Alive | sepsis |
| 3 | 5933 | CGD | HCT | CMV viremia | Deceased | sepsis |
| Defects in intrinsic and innate immunity | | | | | | |
| 1 | 2823 | STAT1 |  |  | Alive | esophagitis, pulmonary |
| 2 | 2955 | STAT1, T cell immunodeficiency, congenital alopecia, and nail dystrophy syndrome | Unspecified |  | Alive | esophagitis |
| 3 | 2982 | STAT1 | Biologic |  | Alive | esophagitis |
| 4 | 3015 | STAT1 |  |  | Alive | esophagitis, CMC |
| 5 | 2872 | STAT1, IgG deficiency |  |  | Alive | esophagitis |
| Not specified | | | | | | |
| 1 | 3291 | Mutation in CFP/CYBB/IKBKG/MAGT1/XIAP | HCT | pseudomonas | Alive | esophagitis |
| 2 | 3558 | DLBCL | Liver transplant, chemotherapy, steroids, calcineurin inhibitor | | Alive | esophagitis |
| 3 | 1055 | DLBCL, neutropenia |  | EBV | Alive | esophagitis |
| 4 | 2933 | Lymphopenia, Trisomy 21 |  |  | Alive | esophagitis |
| 5 | 1387 | Neutropenia | HCT, steroids |  | Alive | esophagitis |
| 6 | 6388 |  | Steroids |  | Alive | esophagitis |
| 7 | 5805 | 5P deletion, ectodermal dysplasia |  |  | Alive | esophagitis |
| 8 | 5605 | Immune disorder |  |  | Alive | esophagitis |
| 9 | 4877 | Abnormal genetic test |  |  | Alive | esophagitis |

Clinical features at time of encounters where patients were diagnosed with *Candidal* esophagitis, laryngitis, sepsis, meningitis, and/or vulvovaginitis. “Steroids” indicates systemic corticosteroid use. “Unspecified” immunosuppression indicates that a patient had a diagnosis code for immunosuppressive medication use but the exact agent was not specified. Abbreviations: Ab: antibody; ADA: adenosine deaminase; ALL: acute lymphoblastic leukemia; ALPS: autoimmune lymphoproliferative syndrome; AML: acute myeloid leukemia; APECED: autoimmune polyendocrinopathy-candidiasis-ectodermal dystrophy; B-ALL: B-cell acute lymphoblastic leukemia; BK: BK polyomavirus; CGD: chronic granulomatous disease; CID: combined immunodeficiency; CFP (complement factor properdin); CMC: chronic mucocutaneous candidiasis; CMV: cytomegalovirus; CTLA4: cytotoxic T-lymphocyte–associated protein 4; CVID: common variable immunodeficiency; CYBB: Cytochrome b-245 Beta Chain; DLBCL: diffuse large B-cell lymphoma; EBV: Epstein–Barr virus; FLG: filaggrin gene; GATA-2: GATA-binding protein 2; HCT: hematopoietic stem cell transplant; HIV: human immunodeficiency virus; HSV: herpes simplex virus; IKBKG: inhibitor of nuclear factor kappa-B kinase regulatory subunit gamma (NEMO); LRBA: lipopolysaccharide-responsive beige-like anchor protein; MAC: Mycobacterium avium complex; MAGT1: Magnesium Transporter 1, MDS: myelodysplastic syndrome; MHC: major histocompatibility complex; NFKB2: nuclear factor kappa B subunit 2; NKX2-1: NK2 homeobox 1; PJP: Pneumocystis jirovecii pneumonia; PTEN: Phosphatase and TENsin homolog, SCID: severe combined immunodeficiency; STAT1: signal transducer and activator of transcription 1; VZV: varicella zoster virus; WHIM: warts, hypogammaglobulinemia, infections, and myelokathexis; XIAP: X-linked inhibitor of apoptosis.
